# From Stability to Complexity: A Systematic Review Protocol on Long-term Divergence Exponents in Gait Analysis

**DOI:** 10.1101/2024.12.02.24318001

**Authors:** Philippe Terrier

**Affiliations:** Haute-Ecole ARC Santé. University of Applied Sciences and Arts Western Switzerland

**Keywords:** Gait analysis, Nonlinear dynamics, Gait disorders, Outcome Assessment, Health Care

## Abstract

Long-term divergence exponents derived from nonlinear gait analysis (maximum Lyapunov exponent method) have recently been reinterpreted as measures of gait complexity rather than stability. This shift necessitates a comprehensive review of existing literature. This systematic review protocol aims to critically examine studies using long-term divergence exponents in gait analysis. The focus will be on reconciling previous findings with current understanding, evaluating methodological approaches, and synthesizing comparable results. We will search Web of Science (including MEDLINE) for peer-reviewed articles published between 2001 and 2024 that report long-term divergence exponents calculated using Rosenstein’s algorithm in human gait studies. Two independent reviewers will screen articles and extract data on study characteristics, methodological specifications, and result interpretations. Primary outcomes will include tracking how result interpretations have evolved over time and identifying potential reinterpretations based on current knowledge. Secondary outcomes will address methodological standardization. Data synthesis will primarily be narrative. Where possible, meta-analyses will be conducted for studies with comparable methods and objectives. Given the expected exploratory nature of many included studies, a narrative assessment of methodological quality will be performed instead of a formal risk of bias evaluation. This review will consolidate understanding of long-term divergence exponents as measures of gait complexity and automaticity, establish standardized computational methods, and inform future research and clinical applications in gait analysis.

## 1 Introduction

The maximum Lyapunov exponent method is a sophisticated mathematical approach, derived from chaos theory, for analyzing nonlinear dynamical systems and their sensitivity to initial conditions [2]. In gait analysis, this method quantifies how the locomotor system responds to small perturbations during walking by examining the rate at which initially adjacent trajectories in state space of gait dynamics diverge over time [3]. The analysis typically involves reconstructing the attractor of the gait dynamics from continuous kinematic or kinetic signals and computing logarithmic divergence curves [4]. Two distinct time scales have emerged as relevant for gait assessment: short-term divergence, computed over the range of 0 to 1 stride, which evaluates the immediate response of the gait to perturbations and is interpreted as a measure of local dynamic stability, and long-term divergence, computed over 4-10 strides, which examines the behavior of the system over multiple gait cycles [5]. While short-term divergence has shown consistent associations with aging gait, fall risk and dynamic balance control [6–8], some studies have revealed puzzling discrepancies, with short-term and long-term divergence often responding differently or even inversely to experimental conditions [9,10], suggesting that they may capture fundamentally different aspects of gait control [11].

While early studies used long-term divergence exponents to assess gait stability [3,12], there is increasing evidence that this measure captures distinct features of gait control than its short-term counterpart. During steady walking, stride parameters fluctuate with complex fractal patterns characterized by long-range correlations between successive strides, reflecting the elaborate organization of locomotor control (gait complexity) [13]. Several studies have shown that long-term divergence exponents are strongly associated with the correlation structure of stride intervals, as measured by scaling exponents that characterize stride-to-stride fluctuation complexity [4,14–17]. Studies comparing normal walking with externally cued conditions found that long-term divergence exponents decreased substantially, while short-term divergence exponents remained largely unchanged [15–17]. These findings replicate what is observed in studies using scaling exponents to characterize gait complexity and cued walking [13]. Further evidence showed that acceleration signals with identical shapes, but modified stride interval complexity exhibited long-term divergence exponents that varied dramatically with the type of correlation structure applied, with differences of up to 66% between anticorrelated and correlated signals [18].

The strong relationship between stride interval complexity and long-term divergence exponents, combined with its weak association with fall risk and stability measures, has led researchers to propose the term “Attractor Complexity Index” (ACI) as a more appropriate name that better reflects their true nature as a measure of gait complexity rather than stability [18]. Recent evidence suggests that gait complexity may reflect the degree of cognitive involvement in locomotor control (gait automaticity [19]), as executive function appears to orchestrate the regulation of stride-to-stride fluctuations [17,20,21]. Therefore, long-term divergence exponents hold potential for advancing clinical gait assessment by providing a more nuanced understanding of gait control, particularly in conditions where traditional measures of stability may fall short. Additionally, the ACI could facilitate the development of wearable technologies for ecological monitoring of gait complexity in real-world settings, offering valuable insights into motor-cognitive interactions during daily activities [17].

This fundamental shift in our understanding of long-term divergence exponents requires a critical re-examination of the existing literature. Many studies published prior to this conceptual evolution interpreted long-term divergence as an index of stability, potentially leading to conclusions that need to be reconsidered in light of its newly recognized role as a measure of gait complexity. While these studies may have misinterpreted their findings through the lens of stability, their data remain valuable and could provide insights when reanalyzed from a new perspective. Moreover, substantial methodological heterogeneity exists across studies, both in the measurement techniques used to capture gait dynamics (including accelerometry, motion capture, and joint angle measurements [22]) and in the algorithmic procedures employed to calculate divergence exponents (such as time delay selection, embedding dimension, and stride range definition).

A systematic review would therefore serve three important purposes: first, to reconcile previous findings with our current understanding of long-term divergence as a measure of gait complexity; second, to evaluate and potentially standardize the experimental and computational methods used across studies to improve the reliability and comparability of future research; and third, to quantitatively synthesize results from comparable studies through meta-analysis when possible.

## 2 Methods

### 2.1 Study Eligibility

The systematic review will include original, peer-reviewed research articles that describe studies of human locomotion, including studies of healthy participants or patients (or both) of all ages. We will consider observational, experimental, and intervention studies, with no restriction on sample size. Studies must use specifically the calculation of divergence curves based on the maximum Lyapunov exponent method using Rosenstein’s algorithm [3,4], with explicit reporting of long-term divergence exponents. We will consider articles published between 2001 and 2024. Studies analyzing running gait or other locomotion modes will be excluded. Similarly, pure modeling studies based on computer simulations of locomotion will not be considered. Studies that do not provide sufficient methodological information to assess the computation of long-term divergence exponents will be excluded, as will review articles, letters, editorials, and conference abstracts (Table 1).

**Table 1.** Inclusion and exclusion criteria.

| Concept | Inclusion | Exclusion |
| --- | --- | --- |
| Type of documents | Peer-reviewed original articles | Review articles, letters, editorials, conference abstracts |
| Publication status | Published | Grey literature |
| Publication date | 2001–2024 | Outside of this date range |
| Language | English or French | Other languages |
| Population | Human studies of all populations | Non-human studies |
| Study design | Observational, experimental, intervention studies | Modeling studies, simulation studies |
| Sample size | All sample sizes (no restriction) | None |
| Locomotion mode | Walking | Running, jumping, other non-walking modes |
| Nonlinear gait analysis | Studies using the maximum Lyapunov exponent method | Other nonlinear methods |
| Lyapunov exponent analysis | Studies using Rosenstein's algorithm, reporting long-term divergence exponents (with or without short-term) | Studies reporting only short-term divergence exponents, studies using other algorithms |
| Methodological reporting | Studies with sufficient methodological details on divergence exponent computation | Studies lacking adequate methodological information on divergence exponent calculation |

### 2.2 Information Sources and Search Strategy

We will search for relevant articles using the Clarivate Web of Science search engine to retrieve references from both MEDLINE and the Web of Science Core Collection databases. The search strategy will be developed using an iterative process, starting with initial keywords based on the research question. We will analyze the text words contained in the titles and abstracts of the retrieved articles, as well as the index terms used to describe these articles, to refine our search terms. If necessary, we will conduct additional searches using newly identified keywords to ensure comprehensive coverage. For example, our preliminary search string could be TS = ((gait* OR walk*) AND (stability OR dynamic* OR nonlinear OR “non-linear”) AND (lyapunov OR “local divergence” OR (divergent exponent*)) AND (child* OR adult* OR people OR subject* OR person* OR participant*)). The complete Web of Science search strategy, including all search terms and Boolean operators, will be provided to ensure reproducibility. While we will include articles in both English and French as these are the languages spoken by the investigators, we anticipate that most, if not all, relevant articles will be in English. The search will include articles published between 2001 and 2024, as 2001 marks the beginning of the application of maximum Lyapunov exponent methods to gait analysis.

### 2.3 Study Selection Process

Two research assistants will independently screen the retrieved articles in two phases. In the first phase, they will review titles and abstracts using broad inclusion criteria, as the specific use of long-term divergence exponents is often not explicitly mentioned in abstracts. This initial screening will likely include studies using various nonlinear methods for gait analysis, including short-term divergence exponents, Wolf’s algorithm [23], and other stability measures. From an estimated initial pool of 200-400 articles, studies that potentially involve nonlinear gait analysis will proceed to the second phase, where their full texts will be thoroughly reviewed to specifically identify those using long-term divergence exponents based on Rosenstein’s algorithm. Additionally, the reference lists of included articles will be hand-searched to identify potentially relevant studies that may have been missed by the electronic database search (Fig.1). Disagreements between the two reviewers will be resolved through discussion, with the principal investigator (PT) acting as arbitrator if consensus cannot be reached. Based on previous systematic reviews in this field [6,22], we estimate that 20-40 articles will meet all inclusion criteria for the outcome analysis.

**Figure 1.**
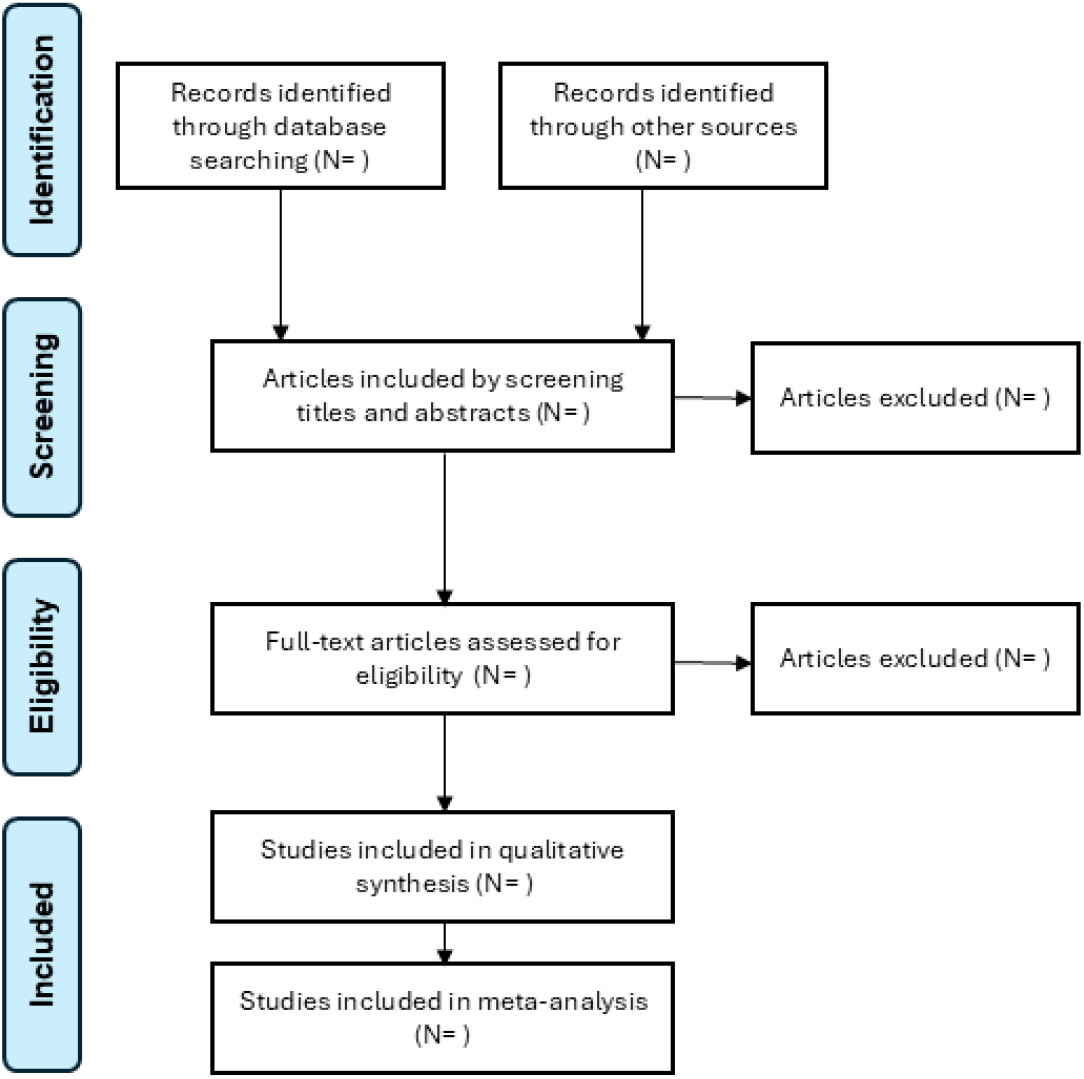
Flow-chart of the selection process.

### 2.4 Data Collection Process and Data Items

The investigators will chart the data from the included studies using standardized extraction tables (Table 2–4). They will collect three main categories of information. First, study characteristics will include the publication year, authors, sample size, participant demographics (age, gender, health status), and research context (methodological study, clinical evaluation, or intervention evaluation). Second, methodological specifications will detail the measurement methods (primary measurement system: accelerometer, goniometer, motion capture; signal type: velocity, acceleration, angle, position; sampling rate; measurement duration and/or number of strides; walking condition: overground or treadmill), signal processing procedures (filtering and resampling methods), and the specific parameters used to compute long-term divergence exponents (time delay, embedding dimension, stride range). Third, we will extract information about the aim of each study, study conclusion, and interpretation of long-term divergence exponent results.

**Table 2.** Data extraction template for study characteristics and population distribution.

| Column label | Description |
| --- | --- |
| Study ID | Unique identifier |
| Authors | Abbreviated author lists |
| Year | Publication year |
| Sample size | Total number of participants |
| Sample composition | Demographic and clinical information |
| Research type | Methodological / clinical / intervention |

**Table 3.** Data extraction template for methodological and technical specifications.

| Column label | Description |
| --- | --- |
| Study ID | Unique identifier |
| Measurement system | Accelerometer, goniometer, motion capture... |
| Signal type | Velocity, acceleration, angle, position... |
| Duration | Stride number or recording duration |
| Walking condition | Overground or treadmill |
| Filtering method | If applicable |
| Resampling method | If applicable |
| Attractor reconstruction | Time delay and embedding dimensions |
| Stride range | Range used for calculation of long-term divergence |

**Table 4.**
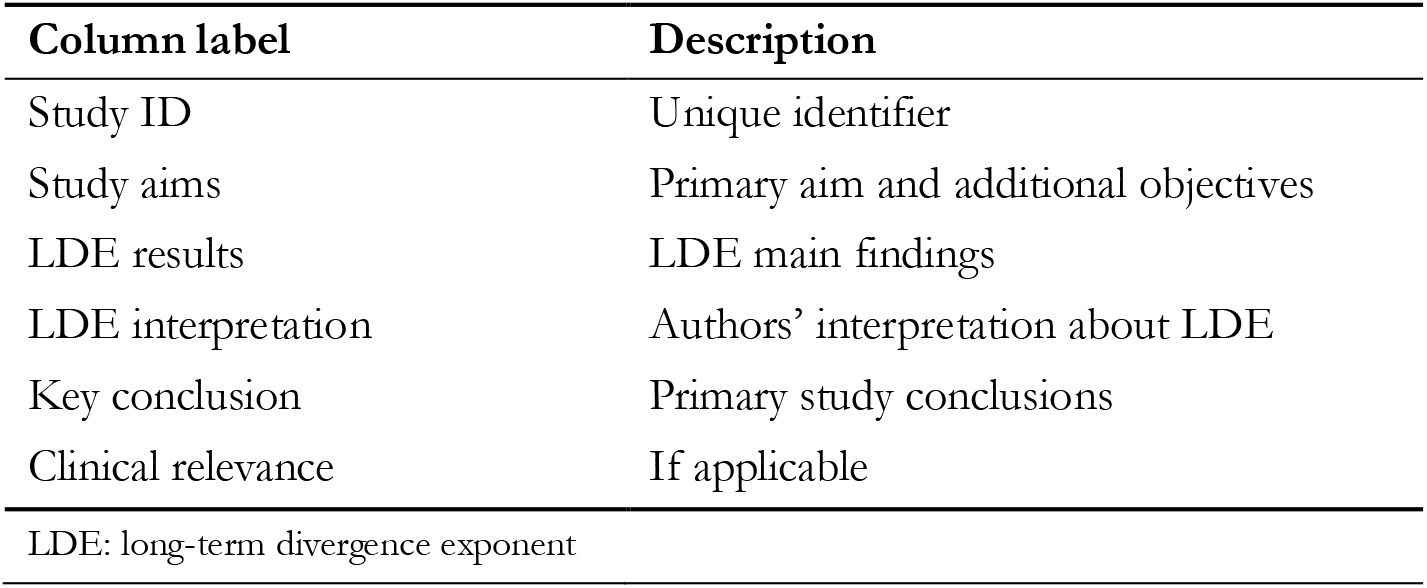
Data extraction template for study aims and long-term divergence exponent findings.

| Column label | Description |
| --- | --- |
| Study ID | Unique identifier |
| Study aims | Primary aim and additional objectives |
| LDE results | LDE main findings |
| LDE interpretation | Authors' interpretation about LDE |
| Key conclusion | Primary study conclusions |
| Clinical relevance | If applicable |
| LDE: long-term divergence exponent |  |

### 2.5 Outcomes and Prioritization

The primary outcomes will focus on findings of the included studies, specifically: how results were interpreted, the evolution of this interpretation over time, and how findings might be reinterpreted in light of current understanding of long-term divergence as a gait complexity and automaticity measure. We will particularly examine relationships between long-term divergence, other gait measures, and experimental conditions that may support its role as a gait complexity index. Secondary outcomes will address methodological standardization, including measurement specifications, signal processing procedures, and computation parameters.

### 2.6 Data Synthesis

The synthesis will primarily be narrative, focusing on the interpretation of long-term divergence exponents and methodological considerations. Additionally, if we identify at least four studies with comparable methods and objectives (e.g., comparing young vs. older adults, or examining specific clinical conditions), we will conduct meta-analyses of effect sizes. For these quantitative analyses, we will calculate standardized mean differences and their 95% confidence intervals. Study heterogeneity will be assessed using I^2^ statistics, and pooled effect sizes will be computed using a fixed-effect model. The results of these meta-analyses will be presented using forest plots.

### 2.7 Risk of Bias Assessment

Given the exploratory nature of many studies using maximum Lyapunov exponents in gait analysis, where the primary aim was often to investigate the potential of the method rather than to test specific hypotheses, a formal risk of bias assessment using standardized tools would not be appropriate. Instead, we will conduct a narrative assessment focusing on three key aspects of methodological quality. First, we will examine analytical rigor, including the completeness of parameter reporting and the appropriateness of computational methods. Second, we will assess the thoroughness of the reporting of results, particularly with respect to the presentation of descriptive statistics and measures of variability. Third, we will discuss sample size considerations, recognizing that many exploratory studies may have used convenience samples rather than formal power calculations. This narrative approach to quality assessment is consistent with the developmental nature of the field, where methodological standardization is still evolving. The results of this assessment will inform recommendations for improving the quality of future research using long-term divergence analysis.

## 3 Dissemination Plans

This protocol has been published as a preprint on medRxiv. The completed systematic review will be submitted for publication in a peer-reviewed journal focusing on biomechanics, gait analysis, or research methodology. The results will be presented at relevant scientific conferences in the field of biomechanics, exercise sciences, or motor control. The results will be disseminated to both the clinical and research communities to promote standardization of long-term divergence analysis methods and proper interpretation of this measure.

## 4 Discussion and Conclusion

Nonlinear analysis of human gait provides unique insights into locomotor control that complement traditional biomechanical approaches [6,24]. While short-term divergence provides information about dynamic stability and fall risks [5,7,8,11,22], long-term divergence (now called the Attractor Complexity Index) is likely associated with gait automaticity and cognitive involvement in locomotor control [15,18]. This knowledge is particularly valuable for assessing the cognitive demands of walking, as reduced gai t automaticity—characterized by greater reliance on attentional resources—has been linked to pathological gait patterns and may serve as a potential biomarker for diagnosing motor-cognitive impairments, monitoring disease progression, and tailoring rehabilitation strategies in clinical populations [19,25–27].

This systematic review aims to consolidate our understanding of long-term divergence as a measure of gait complexity and automaticity and to establish standardized computational methods. The results will inform both research methodology and clinical applications, potentially improving the assessment of gait disorders using wearable technology in unsupervised (free-living) conditions [6,17,24,28]. Standardization of measurement and computational methods would facilitate the implementation of these analyses in both research and clinical settings, leading to more reliable and comparable assessments of gait complexity across populations and conditions.

## Data Availability

No data were collected or analyzed for this protocol. The completed systematic review will include a comprehensive data sharing plan. All data extracted from included studies during the review process will be made available.

## 5 Declarations

### 5.3 Ethics approval and consent to participate

Ethics approval was not required for this systematic review as it involves the analysis of previously published data and does not involve human participants or animal subjects.

### 5.4 Authors’ contributions

PT conceived the study, obtained funding, designed the protocol, and wrote the manuscript. The author read and approved the final version of the manuscript.

## 5.6 Acknowledgements

The author would like to express sincere gratitude to Marco Pedrotti for his support throughout the ACIDS project. Special thanks are extended to colleagues at the institution for their assistance.

## 5.7 Registration

This protocol was not registered in PROSPERO or other systematic review registries due to the exploratory nature of many studies expected to be included and the limited scope for formal risk of bias assessment. Given these constraints, we opted to publish this protocol as a preprint on medRxiv to ensure transparency and allow for peer feedback prior to conducting the full review. Any future amendments to this protocol will be documented and reported in the final systematic review publication.

